## Supplementary figures and images for "Creating a General-Purpose Generative Model for Healthcare Data based on Multiple Clinical Studies"

### S1 Fig

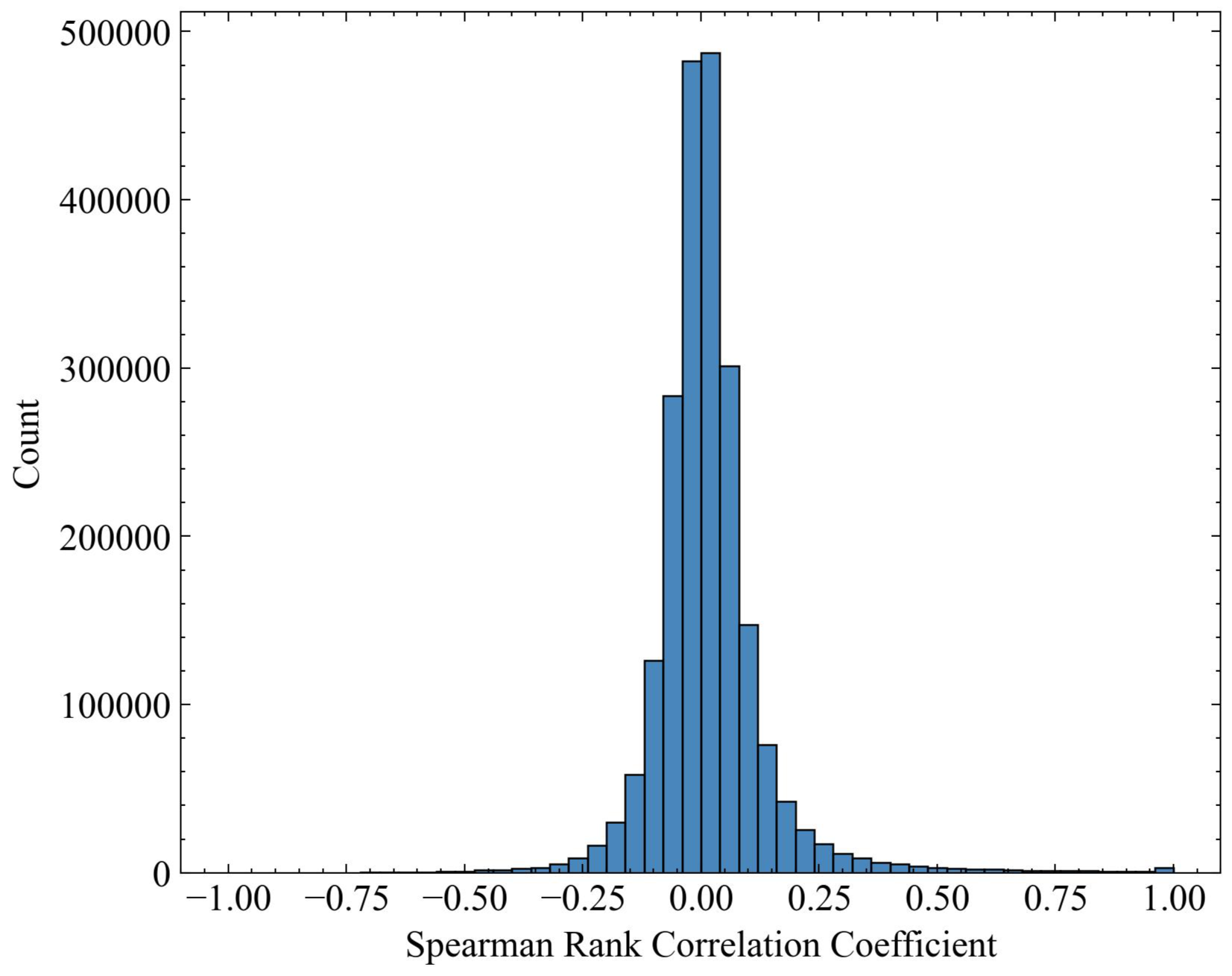

### S2 Fig

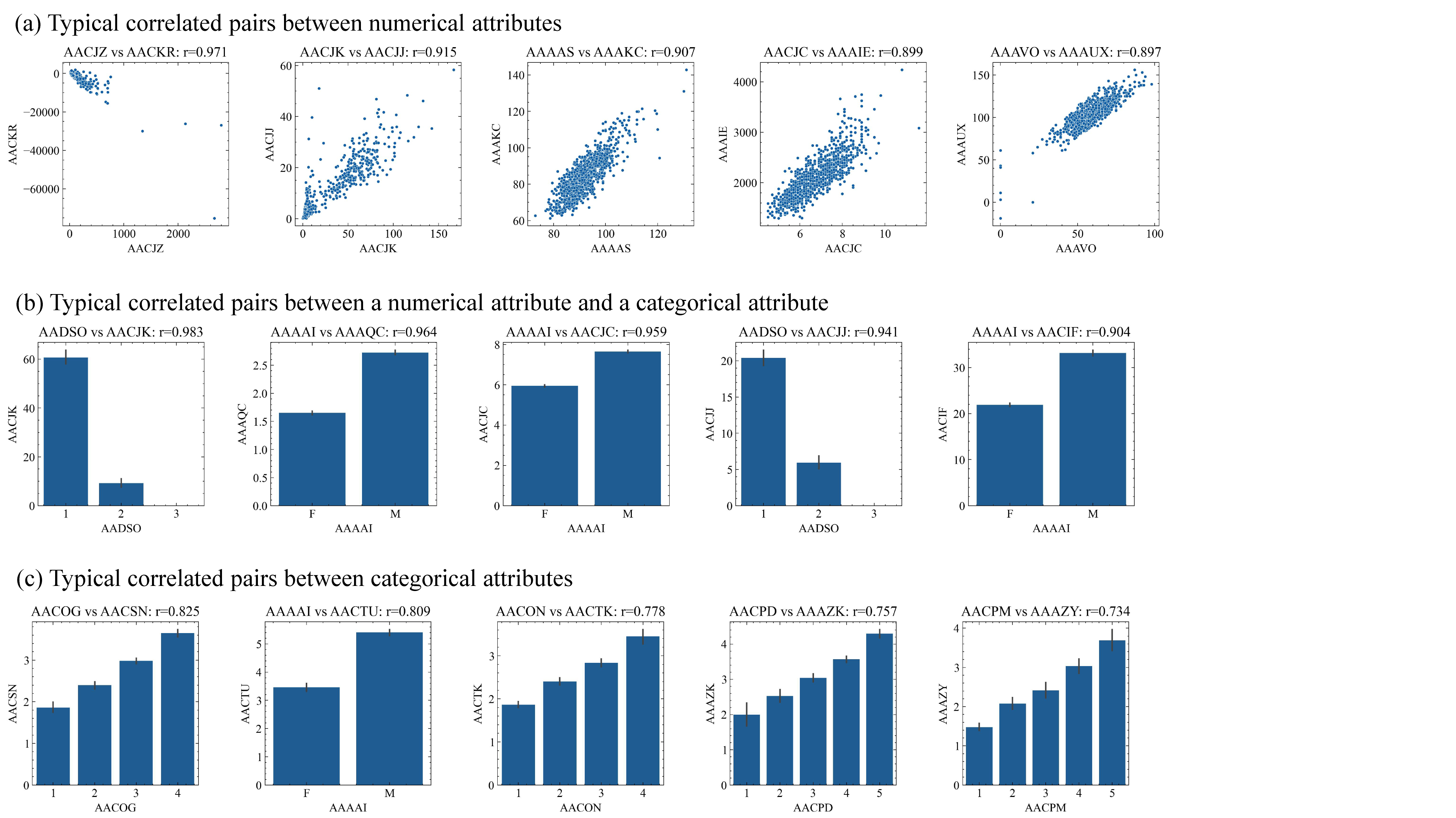
