## Supplementary material for "Creating a General-Purpose Generative Model for Healthcare Data based on Multiple Clinical Studies": S1 Table

**S1 Table Number of male participants.** Decade, N (visit 1), Ratio (\%), N (visit 2), Conversion (%), and Ref. (%) correspond to the age group of participants, numbers in each age group, percentages of each age group relative to the total participants, ratios of N (visit 2) to N (visit 1), and ratios of the Japanese population from the recent Japanese official statistics that are available at e-Stat (2019), respectively. At visit 2, conversion rates of the men’s 60-69 and ≥ 70 age groups were slightly lower compared to those in younger age groups. This reduction appears to be attributable to the exclusion criteria applied in the study or their health conditions (see the medical history records in S3 Table).

| **Decade** | **N (visit 1)** | **Ratio (%)** | **N (visit 2)** | **Conversion (%)** | **Ref. (%)** |
| --- | --- | --- | --- | --- | --- |
| 20-29 | 62 | 6.2 | 52 | 83.9 | 6.2 |
| 30-39 | 73 | 7.3 | 67 | 91.8 | 6.9 |
| 40-49 | 91 | 9.2 | 82 | 90.1 | 8.9 |
| 50-59 | 80 | 8 | 72 | 90 | 9 |
| 60-69 | 78 | 7.8 | 61 | 78.2 | 7.5 |
| ≥ 70 | 100 | 10.1 | 71 | 71 | 10.6 |
