## Supplementary material for "Creating a General-Purpose Generative Model for Healthcare Data based on Multiple Clinical Studies": S2 Table

**S2 Table. Number of female participants.** The column name definitions are the same as S1 Table.

| **Decade** | **N (visit 1)** | **Ratio (%)** | **N (visit 2)** | **Conversion (%)** | **Ref. (%)** |
| --- | --- | --- | --- | --- | --- |
| 20-29 | 61 | 6.1 | 51 | 83.6 | 6 |
| 30-39 | 83 | 8.4 | 73 | 88 | 6.6 |
| 40-49 | 93 | 9.4 | 80 | 86 | 8.6 |
| 50-59 | 84 | 8.5 | 73 | 86.9 | 8.8 |
| 60-69 | 85 | 8.6 | 71 | 83.5 | 7.8 |
| ≥ 70 | 104 | 10.5 | 89 | 85.6 | 13.1 |
