## Supplementary material for "Creating a General-Purpose Generative Model for Healthcare Data based on Multiple Clinical Studies": S3 Table

**S3 Table. Number of diseases under treatment.** Numbers of participants undergoing treatment for each disease.

| **Decade** | **Hypertension**  **(Male)** | **Hypertension**  **(Female)** | **Diabetes**  **(Male)** | **Diabetes**  **(Female)** | **Dyslipidemia**  **(Male)** | **Dyslipidemia**  **(Female)** |
| --- | --- | --- | --- | --- | --- | --- |
| 20-29 | 1 | 0 | 0 | 0 | 0 | 1 |
| 30-39 | 0 | 0 | 1 | 1 | 1 | 1 |
| 40-49 | 8 | 1 | 3 | 0 | 6 | 2 |
| 50-59 | 13 | 2 | 4 | 2 | 7 | 2 |
| 60-69 | 32 | 9 | 9 | 4 | 11 | 9 |
| ≥ 70 | 37 | 15 | 21 | 2 | 19 | 22 |
