## Supplementary material for "Creating a General-Purpose Generative Model for Healthcare Data based on Multiple Clinical Studies": S4 Table

**S4 Table. Attribute number, missing rate, and outlier rate of each measurement.** Measurement categories were described elsewhere [1].

| **Measurement** | **Attribute** | **Record** | **Missing Rate Mean** | **Outlier Rate Mean** |
| --- | --- | --- | --- | --- |
| Blood Pressure and Arterial Stiffness Measurements | 48 | 994 | 0.0052 | 0.006 |
| Lifestyle Investigation and Questionnaire | 1014 | 994 | 0.017 | 0.0114 |
| Cognitive Function Analysis | 69 | 994 | 0.0041 | 0.0138 |
| Laboratory Analysis | 97 | 994 | 0.066 | 0.0095 |
| Oral Glucose Tolerance Test | 57 | 994 | 0.1987 | 0.011 |
| Anthropometric Measurements | 77 | 994 | 0.0021 | 0.0063 |
| Skin Surface Spectroscopy | 9 | 994 | 0.0499 | 0.0057 |
| Physical Performance Tests | 186 | 994 | 0.0687 | 0.0098 |
| Hand Surface Analysis | 4 | 994 | 0.1972 | 0.0088 |
| Liquid Chromatography-Tandem Mass Spectrometry Analysis | 42 | 994 | 0.154 | 0.007 |
| Body Odor Analysis | 19 | 994 | 0.1561 | 0.016 |
| Lipids in the Stratum Corneum and Sebum Analysis | 44 | 994 | 0.0141 | 0.0083 |
| Hair Loss Determination | 17 | 994 | 0.3295 | 0.0085 |
| Lipid Mediator Analysis | 38 | 994 | 0.5726 | 0.0066 |
| SSL-RNA Analysis | 97 | 994 | 0.1433 | 0.0076 |
| Microbiota Analysis | 50 | 994 | 0.0925 | 0.0128 |
