## Supplementary material for "Creating a General-Purpose Generative Model for Healthcare Data based on Multiple Clinical Studies": S1 Text

**SSL-RNA, intestinal microbiota, and saliva microbiota analysis**

1. **Skin surface lipids-RNA**

The skin surface lipids (SSL) collection and SSL-RNA purification were performed as described previously [1]. In brief, SSLs were collected by wiping the whole face (forehead, cheek, face line, nose, and chin) or parietal scalp using an oil-blotting film (8.0 cm × 5.0 cm, 3M Japan, Tokyo, Japan) and samples were stored in glass vials at -80°C until analysis. SSL-RNAs were extracted using QIAzol (QIAGEN, Hilden, Germany). Sequence library preparation was performed using the Ion AmpliSeq Transcriptome Human Gene Expression Kit (Thermo Fisher Scientific). The adapter sequence was ligated to the library and the libraries were eluted from the beads using Library Amp Mix (Thermo Fisher Scientific) containing Library Amp Primers. The quality of the sequence library was assessed using the High-Sensitivity D1000 ScreenTape on the Agilent 4200 TapeStation. An Ion Library TaqMan Quantitation Kit (Thermo Fisher Scientific) was used to quantify the library. All statistical analyses and normalization of the RNA-seq transcriptome data were performed using the R statistical language. Read counts were generated using the AmpliSeq RNA plugin in the Ion Torrent Suite Software (Thermo Fisher Scientific). Among the gene expression analyses, the analysis of 97 genes related to skin function, inflammation, and immunity (see the list below) were normalized by reads per million mapped reads (RPM), and the genes were expressed with a mean of log2 (normalized counts + 1).

**Selected genes:**

*IL1B, STAT1, CCR1, IL8, IRF1, IFNGR1, CCL3, CCL4, IFNGR2, MX1, IL7R, STAT6, IL4R, CCL22, CCL17, CCL20, CXCL2, LCN2, STAT3, PI3, S100A9, S100A8, S100A7, S100P, SERPINB1, AHR, CALML5, IL32, KRT10, SERPINB4, FLG, LCE2D, LCE2C, LCE2A, LCE1F, LCE1D, LCE1C, LCE1B, SOAT1, LPIN1, CLDN1, ANXA5, HMGCS1, ANXA6, CDSN, PSORS1C2, ELOVL5, AWAT1, CLN8, SPTLC1, ELOVL3, FADS2, FADS1, GAL, DHCR7, DGAT2, FAR2, KRT79, ACOT2, DEGS2, CERS3, PPL, KRT23, SPTLC3, ACER1, PNPLA3, CORO1B, CTSH, ELL2, FLNA, GPR183, HCAR2, HECA, HK2, HLA-DPA1, IFITM1, IVL, KRTAP19-1, LTB, MARCKS, MT1X, NFE2L2, PHLDA2, PPP1CB, RAB5A, RHOB, RNASEK, RPL37A, RPS5, SHOC2, TGFB1, TXNIP, ZFP36L2, RAB27A, GPX1, GPX4, SOD1*

2. **Intestinal microbiota**

Intestinal microbiota analysis was performed as previously reported [2,3]. DNA was extracted using an automated DNA isolation system (GENE PREP STAR PI-480 KURABO, Japan). The V3-V4 regions of bacterial and archaeal 16S rDNA were amplified using the Pro341F/Pro805R primers and dual-index method. Barcoded amplicons were paired-end sequenced on 2×301-bp using the MiSeq system with MiSeq Reagent Kit version 3 (600 Cycle) chemistry. The primer sequences on paired-end sequencing reads were trimmed by Cutadapt ver 1.18 with default settings. Paired-end sequencing reads were merged using the fastq-join program with default settings.

Only joined reads with a quality value score ≥ 20 for more than 99% of the sequence were extracted using the FASTX-Toolkit. The chimeric sequences, identified as artifacts of amplification, were detected and removed with the bioinformatics tool USEARCH to ensure data accuracy.

For taxonomy, the Ribosomal Database Project (RDP) Classifier ver 2.13 and TechnoSuruga Lab Microbial Identification database (DB-BA) ver 16.0 (TechnoSuruga Laboratory, Japan) were used. Identification was conducted using the Metagenome@KIN ver 2.2.1 analysis software (World Fusion, Japan), with confidence levels ≥ 0.8 for the RDP Classifier and homology thresholds ≥ 97% and DB-BA database. Bacterial flora were added to the database at both the phylum and genus levels, and the Shannon index for the overall intestinal flora. The analysis focused on 4 major phyla and 30 major genera.

3. **Saliva microbiota**

The saliva microbiota analysis was performed as previously reported [4]. In brief, saliva pellets suspended in sterile water were centrifuged to remove the supernatant and stored at -80°C until analysis. The samples were analyzed by Genome Lead Co., Inc (Takamatsu, Kagawa). DNA was extracted from each sample by both enzyme treatment and zirconia beads (EZ-Beads; Promega). The composition of the saliva microbiota was assessed by high-throughput sequencing of the 16S rRNA gene of the V1-V2 region using the Illumina NovaSeq Platform (Illumina). The 2×KAPA HiFi HotStart ReadyMix (KAPA Biosystems) was used for amplicon polymerase chain reaction (PCR) and index PCR. The PCR products were purified using AMPure XP beads (Beckman Coulter). Sequencing was performed using the NovaSeq 6000 Reagent Kit v1.5 (500 cycles; Illumina) with a 251-bp paired-end sequencing protocol. All raw 16S rRNA sequence data were analyzed using the open-source QIIME2 platform [5], version 2021.2 (qiime2-2021.2). Five species (*Porphyromonas gingivalis, Tannerella forsythia, Treponema denticola, Streptococcus mutans, Streptococcus sobrinus*), 21 genera (*Prevotella*, *Streptococcus*, *Neisseria*, *Veillonella*, *Porphyromonas*, *Alloprevotella*, *Fusobacterium*, *Rothia*, *Haemophilus*, *Schaalia*, *Leptotrichia*, *Saccharibacteria*, *Granulicatella*, *Gemella*, *Capnocytophaga*, *Actinomyces*, *Absconditabacteria*, *Campylobacter*, *Selenomonas,* *Treponema*, *Tannerella*), and 2 α-diversity indices, Chao1 and Shannon, were added to the database to represent the overall saliva microbiota.
