## Supplementary material for "Creating a General-Purpose Generative Model for Healthcare Data based on Multiple Clinical Studies": S2 Text

**The details on the imputation error calculation methods**

We calculated the imputation errors for each data type as follows:

**Real Type:** The absolute difference between the imputed value and the masked value was divided by the estimated standard deviation to obtain the standardized absolute error. The standard deviation was estimated from the unconditional output of the model. The mean value and the standard deviation value of the absolute difference were calculated from all the data sources.

**Positive Type:** Let *Im* be the imputed value and *Ma* be the masked value. Using the distribution parameters of the estimated log-normal distribution, Lognormal(μ, σ^2^), the standardized error based on the absolute difference of the logarithms was calculated using the following formula:

$$\frac{\left| \log_{10} \left( Im+1 \right)-\log_{10} \left( Ma+1 \right) \right|}{\sqrt{\sigma^{2}}/\ln\left( 10 \right)}$$

The distribution parameters were estimated from the unconditional output of the model. The mean value and the standard deviation value of the absolute difference were calculated from all the data sources.

**Count Type:** The absolute difference between the parameter of the estimated Poisson distribution, Po(λ), and the masked value was calculated. The mean value and the standard deviation value of the absolute difference were calculated from all the data sources.

**Categorical Type:** The accuracy was determined by whether the most probable category matched the masked category, and the accuracy was calculated from all the data sources.

**Ordered Categorical Type:** The accuracy was determined by whether the most probable category matched the masked category, and the accuracy was calculated from all the data sources.
