## Supplementary material for "Creating a General-Purpose Generative Model for Healthcare Data based on Multiple Clinical Studies": S3 Text

**Univariate and bivariate analyses methods**

1. **Univariate analysis**

The distributional consistency was evaluated by calculating the overlapping area between the distribution of the training data and the distribution of the unconditional output of the model. Evaluation methods used for each data type are as follows:

**Numerals (Real, Positive, and Count Type)**: The distributional consistency was obtained by calculating the overlapping area of histograms derived from the training data and the model outputs for each attribute in numerals. For the training data histograms, we excluded the top and bottom 0.5% of values and created each histogram with 20 bins (for count type, the number of bins was determined from the minimum and maximum values of the training data), normalizing the frequencies by the total number of records. For the model output histograms, we created each histogram with 20 bins based on the estimated distribution parameters (for count type, the number of bins was determined from the minimum and maximum values of the training data), normalizing each total area to 1 as it represents a probability distribution.

**Categories (Categorical and Ordered Categorical Type)**: The distributional consistency of each attribute in categories was obtained by calculating the overlapping area between the normalized distribution of category values in the training data (normalized to an area of 1) and the probability distribution of category values derived from the model output.

2. **Bivariate analysis**

We evaluated the consistency of conditional distributions between two attributes, *P*(*X*|*Y*) using the following methods, depending on the data type combination:

**Numerals-Numerals/Categories:** For the training data, we excluded the top and bottom 0.5% of values of an attribute for the condition, divided the values into five equal intervals, and then created the other attribute histograms of each interval in the same way as for the univariate analysis. For the model output, the other attribute histograms were created in the same way as for the univariate analysis using estimated conditional distribution parameters that were obtained with central values of the five intervals. The mean of the overlapping areas between the training data and the model output in each interval was used as an approximate measure of the consistency of the conditional distributions.

**Categories-Numerals/Categories**: In the same way as for the univariate analysis, we evaluated the consistencies of the conditional distributions for each categorical value of an attribute under the given condition. The consistency between two attributes was then determined by averaging the overlapping areas of these conditional distributions.
