## Supplementary material for "Creating a General-Purpose Generative Model for Healthcare Data based on Multiple Clinical Studies": S4 Text

**Details on the external dataset validation**

The external validation of the VHGM model was conducted using two independent datasets. This section describes the origin, data collection protocols, and harmonization procedures applied to enable cross-dataset comparison of 24 nutritional intake attributes.

**Training dataset**
The training dataset for the VHGM model comprised dietary intake data collected through a 3-day food consumption record, as described in detail by Hibi et al. (2023). Briefly, participants (N = 997) completed dietary records over multiple consecutive days, with trained dietitians verifying portion sizes and nutrient content. Nutrient intakes (e.g., total energy, macronutrients, vitamins, and minerals) were calculated using the Standard Tables of Food Composition in Japan.

**e-Stat dataset (Japan)**
The first external dataset was obtained from the National Health and Nutrition Survey (NHNS), published by the Ministry of Health, Labour and Welfare of Japan via the Portal Site of Official Statistics of Japan (e-Stat). The NHNS employs a stratified random sampling design, selecting approximately 300 census enumeration areas across Japan. Survey households include residents aged one year and older. Dietary intake data are collected on a single, non-holiday weekday using a one-day weighed household dietary record method. Participants or household representatives record dish names, all ingredients used, their quantities, plate waste, and the proportion consumed by each household member. Trained field workers visit households to provide instructions and confirm recorded details. Nutrient values are calculated using the Standard Tables of Food Composition in Japan. For the purpose of comparison with the VHGM training dataset, only data from participants aged 20 years and older were included in the validation analysis (N = 4,927), ensuring methodological comparability with the training dataset.

**NHANES dataset (United States)**
The second external dataset was the U.S. National Health and Nutrition Examination Survey (NHANES) for the 2017–2020 cycle. NHANES uses a multistage probability sampling design to obtain a nationally representative sample of the civilian, noninstitutionalized U.S. population. Dietary intake is assessed using two nonconsecutive 24-hour dietary recalls administered by trained interviewers. Nutrient values are calculated using the USDA Food and Nutrient Database for Dietary Studies. For comparability with the VHGM training dataset, only data from participants aged 20 years and older were used in the validation analysis, and dietary intake data from the first day of recall only were included (N = 8,544).

**Harmonization of variables**
Across the three datasets, we identified 24 nutritional intake attributes (e.g., total energy, protein, fat, carbohydrate, sodium) that exhibited high semantic and methodological equivalence. Selection criteria included: (1) consistent nutrient definitions, (2) comparable units of measurement, and (3) availability of summary statistics (mean and standard deviation).

**List of the 24 nutritional intake attributes:**

1. Energy (kcal)
2. Protein (g)
3. Carbohydrate (g)
4. Total fat (g)
5. Total saturated fatty acids (g)
6. Total monounsaturated fatty acids (g)
7. Cholesterol (mg)
8. Thiamin (Vitamin B1) (mg)
9. Riboflavin (Vitamin B2) (mg)
10. Niacin (mg)
11. Vitamin B6 (mg)
12. Folic acid (µg)
13. Vitamin B12 (µg)
14. Vitamin C (mg)
15. Vitamin D (D2 + D3) (µg)
16. Vitamin K (µg)
17. Calcium (mg)
18. Phosphorus (mg)
19. Magnesium (mg)
20. Iron (mg)
21. Zinc (mg)
22. Copper (mg)
23. Sodium (mg)
24. Potassium (mg)
